# Advancing Brain Health Equity after Traumatic Brain Injury: A Multi-Stakeholder Global Priority-Setting Study

**DOI:** 10.64898/2026.05.19.26353566

**Authors:** Tatyana Mollayeva, Thaisa Tylinski Sant’Ana, Urooba Shaikh, Rachel Spouge, Sara Hanafy, Esme Fuller-Thomson, Michelle McDonald, Angela Colantonio, Daìthì Cee, Gráinne McGettrick, Brian Lawlor

## Abstract

The impact of social parameters on brain health among people with traumatic brain injury (TBI) has been extensively documented. However, translation of this evidence into policy and clinical practice remains limited. This may reflect a lack of coordinated and equity-driven approaches to brain health that integrate diverse stakeholder perspectives, limiting progress toward equity-oriented research and service delivery models. We conducted a convergent parallel mixed-methods study guided by the REporting guideline for PRIority SEtting of health research (REPRISE). We utilized the PROGRESS-Plus framework (Place of residence, Race/ethnicity, Occupation, Gender/sex, Religion, Education, Socioeconomic status, Social capital, and context-specific parameters) to ensure systematic consideration of social parameters in the study. For Objective 1, we synthesized existing evidence on social parameters and brain health outcomes. For Objective 2, we surveyed people with lived experience of TBI, family members/friends, clinicians, researchers, and community leaders across the globe to assess their prioritization of social parameters relevant to brain health. For Objective 3, we integrated evidence synthesis and stakeholder input through a structured Round Robin consensus activity to prioritize actionable areas for feasibility and impact. The activity culminated in the development of a knowledge mobilization agenda designed to inform equity-centred policy, research, and clinical practice. In Objective 1, we identified 59 publications with evidence on the effect of PROGRESS-Plus parameters on brain health outcomes following TBI. Meta-research highlighted that education, age, and country-level indicators are prognostic for brain health after TBI. In Objective 2, the highest-ranked priorities of 113 stakeholders across four continents (North America, Europe, Africa, and Oceania) were education, access to benefits, and income. These priorities were at the centre of discussion in Objective 3, which comprised idea sharing, refinement and thematic clustering, and a final prioritization poll. The resulting final 15 priorities were organized into two tracks: Track A, actions feasible in the short term, and Track B, longer-term implementation priorities. Building on this priority-setting process, co-created with stakeholders around the globe, the findings provide a roadmap for integration of social parameters in TBI research, knowledge exchange, policy, and practice.

## Introduction

Traumatic brain injury (TBI) is a major cause of death and disability globally (1). It is also a significant source of healthcare expenditure and burden, with indirect costs estimated at $400 billion annually, equivalent to about 0.5% of global economic output (1). This cost does not take into account the burden TBI places on people’s lives due to disrupted roles, responsibilities, and relationships after brain injury (2,3). While TBI affects people of all walks of life, it disproportionately places burden of related healthcare needs and resource utilization on people of advanced age, those affected by socioeconomic deprivation, and those who are exposed to marginalization in the larger society (4–6).

The interactions between TBI and social parameters are complex (7–9). Using the PROGRESS-Plus framework to conceptualize social parameters (i.e., Place of residence, Race/ethnicity/culture/language, Occupation, Gender/sex, Religion, Education, Socioeconomic status, Social capital, and contextual “Plus” parameters such as age and disability), we have previously reported multiple nonlinear associations within and between social parameters and brain health outcomes (7–10). Analysis of participant characteristics in published TBI research highlighted that people who bear disproportional affects of TBI and its adverse outcomes are not those who are most represented in research (10). It has also brought to awareness that participation in research is not simply a personal choice, but is heavily influenced by broader social, structural, and institutional contexts that either facilitate or constrain engagement with research processes (11,12). At the same time, best practices and guidelines in TBI have largely emphasized biomedical and clinical evidence, with comparatively less attention given to social parameters. This constitutes a gap between knowledge and practice.

Advancing patient-centered care necessitates not only filling this gap through integration of social parameters and patients’ perspectives in research, but also considering people’s priorities and what matters most to them (2,3,13,14). This paradigm shift involves complementing the traditional emphasis on clinically relevant predictors of TBI outcomes with a rigorous assessment of patient-valued and family process attributes, to design healthcare services and practices that are effective and aligned with people’s priorities and needs (15).

Stakeholder priority-setting activities are particularly well suited for this purpose (16,17). As a stated-preference method, this approach simulates decision-making by asking respondents to rank social parameters based on what they value the most. This requires respondents to make explicit trade-offs, enabling researchers to quantify the relative importance of each social parameter. In this study, we employed a priority-setting approach to: (1) identify and prioritize evidence gaps regarding the impact of social determinants on brain health outcomes in people with TBI; (2) assess stakeholder perspectives on the relevance and importance of these social factors; and (3) develop actionable recommendations for integrating social determinants into TBI research, healthcare indicators, and policy. Leveraging our global network of fellows and leaders in equity in brain health with interdisciplinary expertise, we aimed to ensure that the identified priorities were both evidence-informed and grounded in stakeholder input. This approach was intended to generate actionable knowledge to support policymakers in making more informed and equitable decisions in TBI care.

## Materials and Methods

### Ethics

This paper reports a multistep Knowledge Synthesis and Mobilization Grant (Brain Health and Reduction of Risk for Age-related Cognitive Impairment - Knowledge Synthesis and Mobilization Grants) funded by the Canadian Institutes of Health Research (CIHR) (18). We obtained research ethics approval from the ethics committees at the clinical institution at which the research took place (University Health Network, UHN REB 24-5397) (S1a File), and the Trinity College Dublin Institute of Neuroscience (S1b File). For more details, please refer to the published protocol (19) and registration within the Open Science Framework (OSF) (20), which also provides access to associated study resources and materials.

### Study Design

We conducted a convergent parallel mixed-methods study guided by the REporting guideline for PRIority SEtting of health research (REPRISE) framework (21). Our first objective was to operationalize a definitions of brain health and PROGRESS-Plus parameters, and conduct a systematic synthesis of the association between PROGRESS-Plus parameters and brain health outcomes in TBI (12,13). Our second objective was to develop, administer, and analyse a structured survey capturing participants’ perceived importance of social determinants in relation to brain health outcomes (22,23). Participants included individuals with TBI, family members/friends, clinicians, researchers, and community advocates around the globe. The third objective was to integrate evidence synthesis findings with stakeholder perspectives using a structured Round Robin consensus methodology involving iterative idea generation, refinement, and ranking. Priorities were evaluated and ranked based on predefined criteria of feasibility and impact. A roadmap of the study methodology is presented in Figure 1.

**Fig 1.**
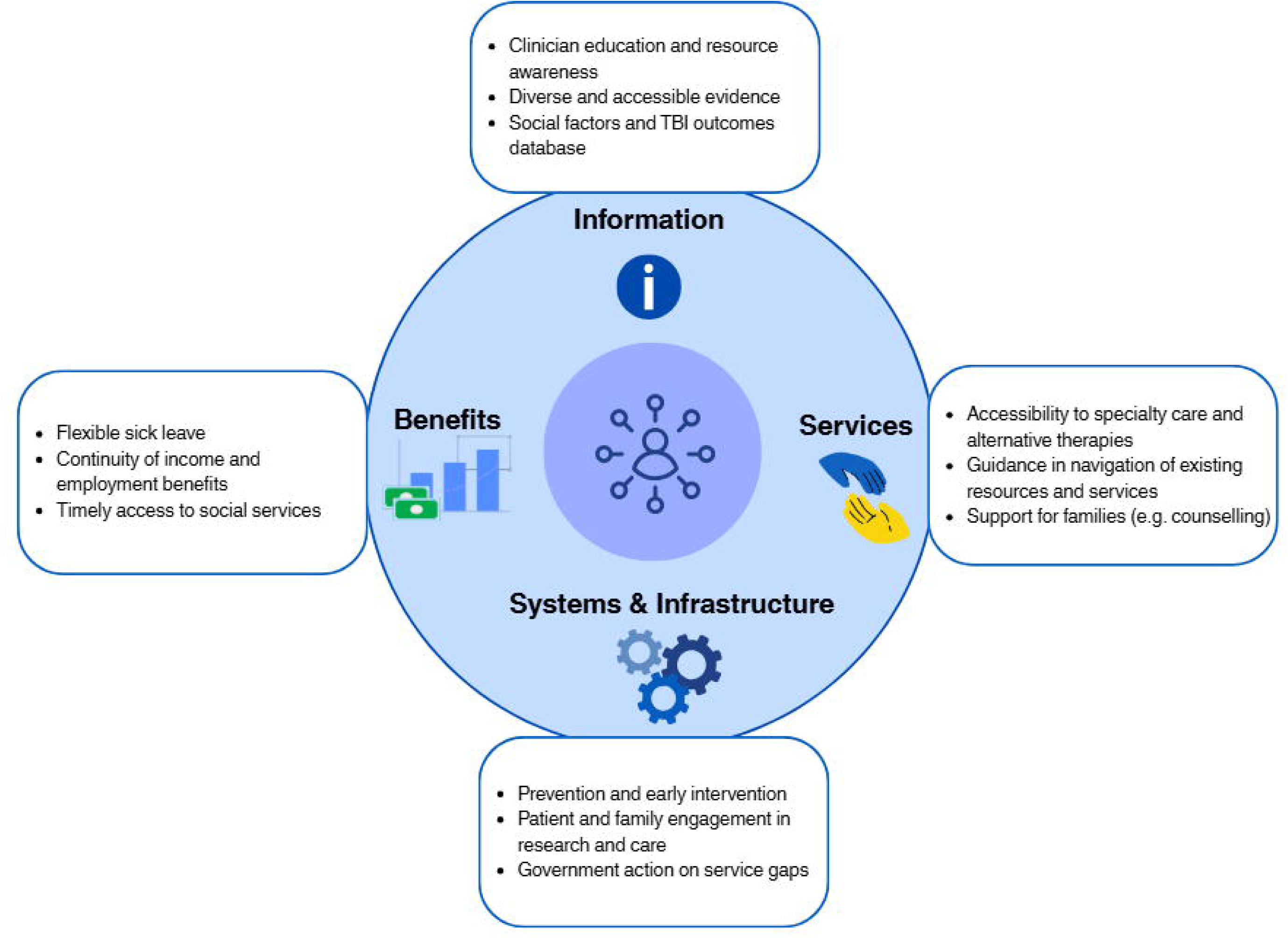
Study design and workflow for each of the three objectives.

### Governance and team

The study was conducted by an interdisciplinary and international team comprising people with lived experience, clinicians, researchers, policy advocates, trainees at multiple levels of training, and representatives from non-governmental organizations (S2 File). Team members who were not on salary support from academic or clinical institutions at the time of the study received an honorarium for their time for participating in meetings and scientific contributions in the amount of $90 per hour, totalling $760 CAD for the duration of the study.

The governance and coordination of the study were led by the principal investigator (TM) and the project manager (TTS). All team members contributed domain-specific expertise and facilitated engagement through their respective professional and community networks, supporting recruitment and stakeholder outreach across patients, family members/friends, clinicians, researchers, and organizational partners.

### Objective 1: Defining brain health and evidence synthesis

#### Establishing Brain Health Definition and Social Parameters

We used a multi-step iterative approach to identify and refine a working definition of brain health and attributes of social parameters.

Published definitions of brain health were extracted from a hybrid concept analysis (9), which synthesized evidence from a literature review and weighted team consensus to select a multidimensional definition of brain health encompassing cognitive, emotional, and motor domains influenced by eco-biopsychosocial determinants (9). The process commenced with team members’ consultation, during which they assessed and rated published definitions of brain health (9); the weighted rankings were then used to define outcome boundaries.

We used the PROGRESS-Plus framework to organize social parameters in data collection, analysis, and reporting. We did not impose a priori limits on PROGRESS-Plus parameters; all relevant parameters were considered eligible if linked to the predefined brain health outcomes (10,15).

#### Evidence Synthesis

To update our published systematic reviews (24,25) on brain health outcomes, with a focus on PROGRESS-Plus parameters, we followed their respective methodologies outlined in detail in the published protocols (26,27).

The data extraction process consisted of three steps. In the first step, we extracted all PROGRESS-Plus parameters from included studies. In the second step, we created a separate matrix for each PROGRESS-Plus parameter. The columns represented each extracted parameter and its measure, and the rows represented the included publications that considered this parameter. In the third step, we identified the frequency ranking of the considered PROGRESS-Plus parameters for each injury severity separately. Next, we completed data transformation and harmonization, following the steps proposed by Kumar et al. (28). We organized results according to the approach taken to investigate the association between PROGRESS-Plus parameters and studied brain health outcomes. In situations where sufficient homogeneity was achieved, we pooled the data for similar outcomes stratified by subgroups of sex/gender and injury severity in meta analysis following methodologies suggested by Cochrane (29). When pooling was not possible, we used analytical approaches to data analysis as suggested by Slavin (30).

As part of the evidence synthesis, we conducted a machine learning (ML) explainable study to identify important PROGRESS Plus parameters relevant to brain health outcomes after TBI, by injury severity (31). To apply ML, we harmonized data on cohorts’ characteristics following the PROGRESS-Plus framework and used the data as predictors of rate of change in cognition after injury (31). We accounted for time from injury to baseline assessment, time between assessments, and country-level structural indicators. We used mean absolute error and root mean squared error to evaluate model performance (32), and Shapley Additive Explanations analysis for explanatory model predictions (33).

### Objective 2: Stakeholder Survey of Priority Preferences

#### Survey development and pilot testing

We extracted data on social parameters reported in included studies and coded them using the PROGRESS-Plus framework. We grouped conceptually similar parameters and transformed them into standardized items, which were mapped to the relevant PROGRESS-Plus domains. We developed the initial list of survey parameters, based on extracted evidence following Cochrane’s recommendations (34), to understand the perspectives and priorities of different stakeholders on the relevance of PROGRESS-Plus parameters to brain health and resources they would find most helpful to support brain health after TBI. This was done by two investigators (TTS and TM).

We shared the preliminary survey with the team for pilot testing and feedback to ensure all relevant items were included in the survey and properly mapped to PROGRESS-Plus domains. We also sought feedback on clarity of the items, comprehensiveness of the descriptor and examples, and any additional relevant items to be included. The final survey, incorporating all feedback, was reviewed and approved by all team members before distribution.

#### Survey distribution

We set a priori target to recruit respondents within five stakeholder groups: (1) persons with lived experience, (2) family/friends, (3) healthcare providers, (4) researchers, and (5) staff/leaders in brain health organizations, who represent a diverse range of gender identities, races/ethnicities, specializations, career stages, professional roles, and affiliations across the globe (19). We restricted the collection of respondents’ location to the continent level to meet ethics requirements on reducing the risk of identifying participants, as the survey was administered through Survey Monkey. We estimated a priori the need to survey at least 10 persons per stakeholder group, requiring at least 50 persons per group (i.e., 10 x 5) (19). Stakeholders could belong to more than one group, and, in that case, they were counted as belonging to the least represented group.

To obtain informed consent, we created a participant information sheet that addressed the purpose and objectives of the research, participant eligibility, risks and benefits of participating, and information about withdrawing from the survey (S3 File). Participants were screened for eligibility according to the following inclusion criteria: being 18 years old or older, able to communicate in English, and self-identifying as belonging to one or more of the following groups: person with TBI; family member or friend who assists a person with TBI with any activity of daily living (i.e., meal preparation, home maintenance, etc.) or provides emotional support without financial compensation; researcher directly involved in TBI research; health care professional who provides care for people with TBI and works with them at least part-time; person holding a staff or leadership position in an organization directly representing or serving people with TBI and their families.

Respondents were asked to review a list of factors related to each PROGRESS-Plus parameter linked to cognitive and brain health outcomes and rank them by perceived level of importance to equity progress using a 5-point scale (1=not important; 5=extremely important). They were also asked what main PROGRESS-Plus parameter research should focus on to support and understand the experiences of people with TBI. Open-ended questions asked participants to explain why they selected their chosen PROGRESS-Plus parameter as a top priority, ideas on how researchers, healthcare workers, and decision-makers could consider social factors to support people with TBI, and to elaborate on helpful information and resources for people with lived experience of TBI and their support system, and the format of delivery for such information.

We used multiple strategies to engage stakeholders in the priority setting process, facilitated by our team members at Brain Injury Canada (35) and the Global Brain Health Institute (36), through their networks with relevant stakeholder organizations. The number and characteristics of the different stakeholders involved enabled assessment of the degree of inclusivity, diversity and equity in the survey. Team members were instructed not to participate in the survey to mitigate the influence of power dynamics, where the perspectives of stakeholders in leadership roles could unintentionally shape or outweigh community responses, and prevent double counting of voices, since many team members were also connected to the organizations and networks through which the survey was distributed. By limiting participation to community respondents, we aimed to ensure that priorities more accurately reflected the perspectives of those the research was intended to serve.

To recruit the target number of participants from each stakeholder group, the survey remained open for five months, from 20/07/2024 to 22/12/2024.

#### Data analysis and weighting priorities

Survey data included quantitative ratings and qualitative input. We integrated the data during analysis by applying descriptive statistical methods to closed-ended responses and qualitative analysis to open-ended responses. We scored each priority domain based on respondents’ ratings, and calculated summary statistics, including frequency distributions, means, and weighted scores to identify the relative importance assigned to each PROGRESS-Plus parameter, overall and by each stakeholder group. To generate an overall priority ranking, we aggregated responses using a weighted scoring approach. For example, a rank-order weighting method (i.e., inverse ranking) was applied so that higher-ranked items contributed more heavily to the final score. We also examined results by stakeholder group, respondents’ continent, race, sex at birth, and age range to assess variation in priority patterns.

We analysed open-ended responses using thematic analysis. First, two researchers (TTS and TM) reviewed the responses together and inductively coded them to identify recurring concepts, concerns, and suggested priorities. Then, the same two researchers grouped coded responses into broader thematic categories that aligned with the predefined PROGRESS-Plus domains. To strengthen analytical rigour, we followed an iterative process to refine themes through several rounds of discussions. Analytical decisions were documented with linking each individual quote to themes to strengthen transparency and replicability.

We visualised priority topics on PROGRESS-Plus domains and resource types, and their weighted scores overall and stratified by stakeholder group, allowing comparison of prioritization patterns across groups, where quantitative weighting reflected relative importance assigned by respondents within each group. We integrated qualitative responses descriptively to provide contextual insight and support interpretation of the quantitative findings. Where qualitative data revealed additional nuance to the quantitative data, we featured these insights for consideration and discussion during Round Robin activity.

### Objective 3: Integrated Round Robin Consensus Process for Priority Setting

#### Pre-Round Robin activity

A summary of the results of the priority setting survey (Objective 2) was shared with the research team. We asked each team member to review results and rate their top two PROGRESS-Plus parameters and top two resource types arising from the priority-setting survey to focus on for knowledge translation, considering both feasibility and potential impact. They were also asked to prepare ideas for two knowledge translation tools and/or mobilization strategies that focus on the top PROGRESS-Plus parameter(s) identified by external stakeholders. We asked them to complete the survey a few days prior to the Round Robin activity to allow for the Study Coordinator and PI to compile them into a single file that was shared with team members to refer to the day before the activity. The procedure is described elsewhere (19).

#### Round Robin activity procedure

We conducted a structured, three-round Round Robin activity via videoconferencing (i.e., Zoom) to facilitate team discussion and prioritize selection of knowledge translation ideas, grounded in stakeholder preferences, feasibility, and potential impact. In Round 1, the facilitator (TTS) asked each team member to present their pre-submitted priorities to the team. As each member shared, other members were asked to provide constructive feedback and build on the ideas. This iterative exchange aimed to deepen understanding, clarify concepts, and raise considerations related to feasibility and impact. The PI (TM) and the project manager (TTS) then collaboratively synthesized the discussion by identifying recurring themes, promising concepts, and gaps requiring further exploration, and shared them with team members prior to Round 2. Round 2 centered on collective organization of ideas into coherent categories to streamline subsequent prioritization.

After the first two rounds, team members were given one week to review detailed summaries of the discussions and thematic analyses from Round 1 and Round 2. This reflective period enabled team members to reconsider and refine their perspectives before the final voting. In Round 3, team members completed a structured electronic poll to rank the consolidated ideas across two predefined tracks: Track A, feasible within the current grant, and Track B, ideas for future grants’ development, with a lower numbered rank indicating higher priority. This voting process generated a final ranked list of the top three priorities per track, based on collective consensus.

#### Evaluation and feedback

Our approach for evaluating the priority setting process was centered on pre-defined criteria of acceptability, replicability, reliability and usefulness informed by the REPRISE framework and related methodological literature (21,30). Evaluation was based on observational data, including level of participation and stakeholder engagement by group, quality of responses to open-ended questions, and the discussion and feedback provided during all stages of the priority setting process. Usefulness was assessed through evaluation of engagement metrics for knowledge translation products through repository analytics. We emphasized transparency and reproducibility by positioning all research-related materials and outputs in an open-access repository (20) and supplementary material for replication by external researchers.

#### Implementation and knowledge mobilization

We followed a structured knowledge mobilization plan that was set a priori in the study protocol (19). Strategies within Tracks A and B were translated into actionable outputs. We engaged trainees and volunteers in the development of knowledge translation products, including evidence synthesis summaries and stakeholder-appropriate material to ensure accessibility and relevance to diverse audiences. We produced a report in plain language on each stage of the priority setting process with tailored summaries for each objective, and made the material available through open access dissemination. We also shared it directly with the leadership of key organizations involved in brain injury and brain health, including the Canadian Institutes of Health Research (CIHR) (37), Brain Injury Canada (35), Acquired Brain Injury Ireland (38), and the Global Brain Health Institute (36), to ensure that findings reached decision-makers who influence policy, funding, and research priorities. Finally, we provided national and international brain injury and brain health organizations with links to publications and emerging knowledge mobilization products, facilitating broader access to the outputs beyond the immediate research team. To maximize accessibility and global reach, our bilingual team members translated key findings from English to French as part of Track A activities.

## Results

### Objective 1. Defining brain health and relevant social parameters

In Objective 1, the working definition of brain health prioritized by the team was “a state of brain function across cognitive, sensory, socio-emotional, behavioural, and motor domains (39).” The poll and results of the weighting process are presented in S4 File. Social parameters relevant to brain health, structured using the PROGRESS-Plus framework (34), can be accessed in S5 File. In parallel, we updated published systematic reviews (40,41) and extracted the social parameters relevant to brain health using the PROGRESS-Plus framework (S5 File). For a systematic review examining the association between PROGRESS-Plus parameters and cognitive outcomes, 28 studies were included comprising 2,407 participants (mean age 32.8 years; 75.5% male) with moderate-to-severe TBI ((20). Only half of the included studies accounted for at least one PROGRESS-Plus parameter in their analyses, most commonly age, sex/gender, and education. Studies that did not adjust for any PROGRESS-Plus parameters more frequently reported cognitive improvement compared to those that accounted for one or more parameters (41). For a systematic review on functional outcomes and life satisfaction, 30 studies were included, representing 101,239 participants with TBI (mean age 38.8 ± 17.1 years; 85% male) across injury severities (40). All studies included at least one PROGRESS-Plus parameter in their analyses, with age, sex/gender, social capital, education, and occupation most frequently reported. Among the eight studies (28%) that built multivariate models, race was associated with life satisfaction, and race, language, education, and marital status were associated with functional outcomes (40).

To further explore the relevance of social parameters for brain health outcomes after TBI, we utilized ML algorithms (i.e., random forest, gradient boosting, and extreme gradient boosting) to data extracted from 30 published studies including over 2,000 participants with TBI (31). Results highlighted country-level structural indicators (Gender Inequality Index, GII) (42), age, and variation in education as key PROGRESS-Plus predictors for rate of change for both injury severities (31). Other PROGRESS-Plus characteristics of study charts (i.e., race, occupation, socioeconomic status, social capital) were limited in reporting (<25% coverage), resulting in their exclusion from the analytical models. Sensitivity analyses for predicting rate of change in executive function and learning and memory confirmed the robustness of the results on the importance of variation in education, age, and GII of country of cohort origin as predictors of cognitive outcome after TBI (31). These parameters were identified as important for consideration within the feasibility and impact assessment during the priority setting. Taking into account the gaps in existing evidence, survey development and the following objective were intentionally designed to remain broad, enabling comprehensive capture of relevant social equity determinants without restriction.

### Objective 2: Stakeholder survey of priority preferences

#### Priority setting survey development and pilot-testing

The initial survey and its final version are presented in S6 and S7 Files. Team members highlighted a number of concerns in the initial version, including the need to improve visuals, simplify language, and reduce potentially identifiable information to the few most relevant (i.e., stakeholder group, age range, sex at birth, gender, race, occupation, continent of residence, and whether they live in a rural or urban/suburban area). In response, we modified the survey and assessed its readability using the Flesch Reading Ease formula (43,44), aiming for scores of 60 or higher since the range of 60 to 70 is equivalent to Grade 7/8 reading level (43,45). We employed principles of scientific communication and graphic design, such as alignment, hierarchy and repetition, and accessibility to develop visuals that facilitate comprehension of the written content (14). The final survey was pilot tested among team members before distribution through the online Survey Monkey platform.

#### Survey results

One hundred and thirteen people responded to the survey, and their characteristics are presented in Table 1. Of these, 62.8% were people with TBI, 18.6% were family or friends of people with TBI, 10.6% were researchers, 11.5% were healthcare professionals, and 10.6% were staff/leaders from community organizations serving people with TBI. The age range represented by the greatest number of participants was 51 to 60 years (29.2%). The majority of participants were of female sex (72.6%) and identified as women (71.7%). In terms of race, most participants self-identified as White (85.8%). The majority resided in North America (88.5%) and described their place of residence as an urban city or large town (70%). For occupation, almost half of respondents selected their current work status as employed (44.3%). When asked how long they had been living with, supporting a person with, or working in the field of TBI the most commonly selected response was over 10 years (42.5%).

**Table 1.** Number of respondents per group and their characteristics.

| Respondents' characteristics | Total | Person with TBI | Family member/friend of person | Researcher's working in the field of TBI | Healthcare professional dealing with TBI | Staff leadership in TBI organization | More than one designation* |
| --- | --- | --- | --- | --- | --- | --- | --- |
|  |  |  | with TBI |  |  | on |  |
| **Number** | 113 | 66 | 15 | 5 | 7 | 6 | 14 |
| **Age** |  |  |  |  |  |  |  |
| 18-30 (%) | 8.8 | 4.5 | 6.7 | 40.0 | 14.3 | 16.7 | 14.3 |
| 31-40 (%) | 18.6 | 15.2 | 6.7 | 40.0 | 28.6 | 50.0 | 21.4 |
| 41-50 (%) | 18.6 | 19.7 | 13.3 | 20.0 | 28.6 | 33.3 | 21.4 |
| 51-60 (%) | 29.2 | 37.9 | 13.3 | 0.0 | 28.6 | 0.0 | 14.3 |
| 60+ (%) | 24.8 | 22.7 | 60.0 | 0.0 | 0.0 | 0.0 | 28.6 |
| **Sex at birth (%)** |  |  |  |  |  |  |  |
| Female | 72.6 | 63.6 | 80.0 | 80.0 | 85.7 | 100.0 | 85.7 |
| Male | 27.4 | 36.4 | 20.0 | 20.0 | 14.3 | 0.0 | 14.3 |
| Other | 0.0 | 0.0 | 0.0 | 0.0 | 0.0 | 0.0 | 0.0 |
| **Gender (%)** |  |  |  |  |  |  |  |
| Women | 71.7 | 63.6 | 73.3 | 80.0 | 85.7 | 100.0 | 85.7 |
| Men | 27.4 | 36.4 | 20.0 | 20.0 | 14.3 | 0.0 | 14.3 |
| Non-binary | 0.0 | 0.0 | 0.0 | 0.0 | 0.0 | 0.0 | 0.0 |
| Other/prefer not to say | 0.9 | 0.0 | 6.7 | 0.0 | 0.0 | 0.0 | 0.0 |
| **Race (%)** |  |  |  |  |  |  |  |
| Black | 6.2 | 7.6 | 6.7 | 0.0 | 0.0 | 0.0 | 7.1 |
| East Asian | 0.9 | 1.5 | 0.0 | 0.0 | 0.0 | 0.0 | 0.0 |
| Hispanic or Latino | 1.8 | 1.5 | 6.7 | 0.0 | 0.0 | 0.0 | 0.0 |
| Indigenous | 4.4 | 4.5 | 6.7 | 0.0 | 0.0 | 0.0 | 7.1 |
| Middle Eastern | 0.9 | 0.0 | 0.0 | 0.0 | 0.0 | 16.7 | 0.0 |
| Southeast Asian | 3.5 | 1.5 | 0.0 | 40.0 | 14.3 | 0.0 | 0.0 |
| White | 85.8 | 89.4 | 80.0 | 40.0 | 85.7 | 83.3 | 92.9 |
| Other/multiple/unknown/prefer not to say** | 1.7** | 1.5** | 6.7** | 20.0 | 0.0 | 0.0 | 0.0** |
| **Continent of living (%)** |  |  |  |  |  |  |  |
| Africa | 1.8 | 1.5 | 0.0 | 0.0 | 0.0 | 0.0 | 7.1 |
| Antarctica | 0.0 | 0.0 | 0.0 | 0.0 | 0.0 | 0.0 | 0.0 |
| Asia | 0.0 | 0.0 | 0.0 | 0.0 | 0.0 | 0.0 | 0.0 |
| Europe | 6.2 | 0.0 | 6.7 | 0.0 | 42.9 | 33.3 | 7.1 |
| North America | 88.5 | 97.0 | 93.3 | 60.0 | 57.1 | 66.7 | 78.6 |
| Oceania | 3.5 | 1.5 | 0.0 | 40.0 | 0.0 | 0.0 | 7.1 |
| South America | 0.0 | 0.0 | 0.0 | 0.0 | 0.0 | 0.0 | 0.0 |

| **Area of living (%)** |  |  |  |  |  |  |  |
| --- | --- | --- | --- | --- | --- | --- | --- |
| City or large town (urban) | 61.9 | 59.1 | 66.7 | 100.0 | 71.4 | 50.0 | 57.1 |
| Residential area near a city or large town | 16.8 | 15.2 | 13.3 | 0.0 | 0.0 | 50.0 | 28.6 |
| Countryside (rural) | 18.6 | 22.7 | 20.0 | 0.0 | 28.6 | 0.0 | 7.1 |
| Other/unknown<br>prefer not to say | 2.7 | 3.0 | 0.0 | 0.0 | 0.0 | 0.0 | 7.1 |
| **Current work status (%)** |  |  |  |  |  |  |  |
| Employed | 44.2 | 22.7 | 33.3 | 100.0 | 100.0 | 100.0 | 85.7 |
| Unemployed | 13.3 | 22.7 | 0.0 | 0.0 | 0.0 | 0.0 | 0.0 |
| Retired | 21.2 | 22.7 | 60.0 | 0.0 | 0.0 | 0.0 | 0.0 |
| Other/unknown<br>Prefer not to say | 21.2 | 31.8 | 6.7 | 0.0 | 0.0 | 0.0 | 14.3 |
| **Years living with/attending to someone with/working in the field of TBI (%)** |  |  |  |  |  |  |  |
| <1 year | 5.3 | 6.1 | 0.0 | 20.0 | 14.3 | 0.0 | 0.0 |
| 1-5 year | 27.4 | 27.3 | 40.0 | 40.0 | 14.3 | 0.0 | 28.6 |
| >5-10 years | 24.8 | 27.3 | 13.3 | 20.0 | 28.6 | 33.3 | 21.4 |
| 10+years | 42.5 | 39.4 | 46.7 | 20.0 | 42.9 | 66.7 | 50.0 |

\*Multiple categories includes research participants who selected more than one group. Each of these groups exceeded the minimum requirement of 10 participants according to the protocol.
\*\*The race % sums >100 because several stakeholders chose to identify with more than one race category.

Across all stakeholder groups, social capital was most frequently selected as the highest priority PROGRESS-Plus parameter (20.3%), followed by socioeconomic status (17.7%) and place of residence (14.2%). Less respondents prioritized plus parameters (e.g., age, disability, sexual orientation, etc.; 10.6%), occupation (9.7%), gender and/or sex (7.1%), and education (4.4%). Fewest respondents selected race (2.6%) and religion (1.8%) as their top priority.

By stakeholder group, the top three PROGRESS-Plus parameters selected as top priority by people with TBI were social capital (24.2%), place of residence (16.7%), and socioeconomic status (13.6%). Family/friends of people with TBI selected place of residence (26.7%), education (20.0%), social capital, and socioeconomic status (13.3% each). Healthcare providers prioritized socioeconomic status (57.1%) and gender/sex, occupation, and social capital (14.3% each); staff of TBI organizations prioritized social capital (33.3%) and education, gender/sex, plus, and socioeconomic status (16.7% each), and researchers prioritized socioeconomic status (40%) and gender/sex, place of residence, and plus parameters (e.g., age, (dis)ability, sexual orientation, etc.) (20% each) (S8 File). Results stratified by age range, race, sex at birth, and continent of residence are presented in S8 File.

#### Data analysis and weighting priorities

The weighted results by stakeholder group and overall are presented in Table 2. The highest scored factors across stakeholder groups were access to benefits (average score of 3.75), homelessness (average score of 3.74), and disability (average score of 3.68).

**Table 2.** Weighted priority scores by stakeholder group and overall for PROGRESS-Plus factors. The scores were calculated by weighting group-specific ratings, taking into account the number of respondents in each group. Dash (-) indicates that the factor was not prioritized by respondents within that particular group. Bolded values indicate overall weighted priority scores.

| PROGRESS-Plus domain | Factor | Pers on with TBI | Family/friend | Staff of TBI organization | Healthcare providers | Researcher | Multiple roles | Overall |
| --- | --- | --- | --- | --- | --- | --- | --- | --- |
| Place of residence | Housing insecurity | 3.67 | 3.86 | 3.83 | - | 4.00 | - | - |
|  | Homelessness | 3.67 | - | - | 4.00 | 4.00 | 3.86 | **3.74** |
| Race/ethnicity/culture/language | Language | 3.06 | 3.31 | - | 2.67 | 3.60 | 3.38 | **3.16** |
|  | Race | - | - | 3.50 | - | - | - | - |
| Occupation | Access to benefits | 3.77 | 3.79 | 3.40 | 3.57 | 4.00 | 3.71 | **3.75** |
|  | Work status | - | - | - | 2.57 | - | - | - |
|  | Workplace health & safety | - | - | 3.40 | - | - | - | - |
| Gender/sex | Gender | 2.89 | - | 3.60 | - | 3.60 | - | - |
|  | Sex at birth | - | 3.00 | - | 2.60 | 3.60 | 3.38 | **3.08** |
| Religion | Spirituality | 2.79 | - | 2.67 | 2.50 | 2.40 | - | **2.77** |
|  | Religious beliefs | - | 3.33 | - | - | - | 3.10 | - |
|  | Irreligion | - | - | - | - | - | - | - |
| Education | Level of formal education | 2.98 | 3.21 | 3.00 | 3.50 | 3.40 | 3.38 | **3.12** |
| Socioeconomic status | Income level | 3.30 | 3.50 | 3.60 | 3.14 | 3.40 | 3.57 | **3.37** |
|  | Wealth and assets | - | - | - | - | 3.40 | - | - |
| Social capital | Trust in relationships | 3.63 | 3.67 | - | - | - | - | **3.56** |
|  | Community participation | - | - | 4.00 | 3.29 | 3.60 | - | - |
|  | Social network size | - | - | - | - | - | 3.57 | - |
| Plus | (Dis)ability | 3.66 | 3.79 | - | 3.71 | 4.00 | - | **3.68** |
|  | Exposure to assault/abuse | - | - | - | - | 4.00 | 3.71 | - |
|  | Age | - | - | 3.50 | - | - | - | - |

The top three priorities for people with TBI were access to benefits (3.77), housing insecurity and homelessness (3.67 each), and disability (3.66). For family and friends, the top three scores were for housing insecurity (3.86), access to benefits and disability (tied for second highest weighted priority with score of 3.79 each), and trust in relationships (3.67).

Staff of TBI organizations’ highest scores were for community participation (4.00), gender and income level (3.60 each), and race and age (3.50 each).

Healthcare providers’ top scores were for homelessness (4.00), disability (3.71), and access to benefits and work status (tied for third highest weighted priority with score of 3.57 each).

For researchers, multiple priorities received the same weighted scores and for first, second, and third highest rankings. They were housing insecurity, homelessness, access to benefits, disability, and exposure to assault/abuse as highest (4.00 each), followed by language, gender, sex at birth, and community participation (3.60 each), and level of formal education, income level, and wealth and assets (3.40 each).

People who identified with more than one stakeholder group’s highest scores were for homelessness (3.86), exposure to assault/abuse and access to benefits (tied for second highest weighted priority with score of 3.71 each), and income level and social network size (tied for third highest weighted priority with score of 3.57 each).

Following thematic analysis of responses to the open-ended question, “What resources would help you better understand the links between brain health and social factors? Your response can refer to resources for people with brain injury, their families, and professionals in the field,” four main categories of resources emerged: (**1**) *Information-based Resources*, including training and resources for clinicians, accessible research, brain health information, education, lived experience perspectives, and information on the links between PROGRESS-Plus parameters and TBI; (**2**) *Service-based Resources*, including access to care, alternative treatments, family counselling, resource facilitation, social services, support for challenging behaviours, and telehealth; (**3**) *Systems and Infrastructure*, including assisted transportation, collaboration with people with lived experience in research, family/friend involvement in care, government assessment of service gaps, partnership with organizations, and prevention; and (**4**) *Benefits*, including provision of sick leave. Additionally, some respondents provided specific recommendations for existing resources, such as online programs, webinars, and resources from brain injury organizations. Figure 2 depicts these categories, and each category linked to specific participant responses can be accessed in S9 File.

**Fig 2.** Survey-based themes and content areas for discussion during Round Robin Activity.

The goal is to provide access and link for people with lived experience to resources across the continuum of care and reducing accessibility barriers. The content areas that were discussed at the Round Robin activity for feasibility and impact, listed in the boxes, are not exhaustive. Complete list of topics is listed in S9 File.

### Objective 3: Round Robin consensus process for priority setting

#### Pre-Round Robin activity

We emphasized the Feasibility, Impact, Development, and Execution (FIDE) principles to structure analysis of team member input, informed by established implementation science and knowledge translation frameworks (46,47). Ten core team members (S10 File) completed a survey prior to the Round Robin activity and selected the top two knowledge translation categories they believed to be most important and feasible to address. The breakdown of theme selection by team members was as follows: Information and Services were the most frequently selected themes (eight each) followed by Systems (four) and Benefits (two). Two team members did not complete the survey prior to the meeting, they participated in the Round Robin activity and verbally identified their top categories (S11 File).

#### Round Robin activity

The Round Robin activity consisted of three rounds (S12 File).

In Round 1 of the activity, team members discussed their pre-survey selected priorities. Of the eight stakeholders that selected Information as one of their top two resource types in the pre-survey, six discussed this theme during the Round Robin activity. Of the eight team members that selected Services, three discussed this theme, and of the four that selected Systems, three discussed this theme during the Round Robin. Of the two stakeholders that selected Benefits as one of their top two resource types, neither discussed this theme during the Round Robin. The percentage of pre-selected themes that were not discussed in the Round Robin were 25%, 63%, 25%, and 100% for Information, Services, Systems and Infrastructure, and Benefits, respectively. Therefore, Services and Benefits were discussed the least (S11 File). Fifteen knowledge mobilization ideas emerged from the Round 1 discussion, falling under seven content areas as categorized by the PI and Study Coordinator: partnerships, centralized resource/services database, arts-based/creative engagement, clinician education, marketing and creative messaging, integrated models of care, and ideas for research and future grants.

In Round 2, team members discussed feasibility, impact, and alignment with survey results, and refined content areas through suggested modifications, improvements, or integrations (S11 File). In total, the seven content areas, Partnerships, Centralized Resources/Services Database, Clinician Education, Arts-based/Creative Engagement, Integrated Models of Care, Ideas for Research and Future Grants, and Marketing and Creative Messaging were discussed 17 times across stakeholders. Of the 17 instances, four discussions incorporated all FIDE principles, a study-defined framework informed by established implementation and knowledge translation approaches (23,24). The content areas that reflected all FIDE principles included partnerships, a centralized resources/services database, integrated models of care, and research and future grants. Overall, nine of the 17 discussions incorporated three FIDE principles. Of the nine discussions containing three FIDE principles, Feasibility was discussed seven times, Impact was discussed nine times, Development (further) was discussed three times, and Execution (implementation) was discussed eight times. Out of the 17 discussions, four incorporated two FIDE principles with Feasibility being discussed three times, Impact being discussed one time, Development (further) not being discussed, and Execution (implementation) being discussed three times. All team members referenced at least one FIDE principle in their contributions (S11 File).

Following the first two rounds, team members conducted an interim review of the proposed knowledge mobilization ideas (S13 File). For Track A (feasible within the current grant), the top three priorities ranked by team members were: (1) engaging with federal agencies to mobilize knowledge generated from this research at the national level; (2) promoting existing brain injury service directories, including those developed by Brain Injury Canada (35) and the ABI Lab (48), and searching for additional services to be added to such directories; and (3) integrating existing service directories into patient portals to facilitate accessibility (Table 3). For Track B (future grant opportunities), the top three priorities were: (1) strengthening connections with community partners through Brain Injury Canada and Acquired Brain Injury Ireland (38) to foster long-term relationships and co-develop resources tailored to community needs; (2) integrating existing training programs for healthcare professionals into continuing education frameworks; and (3) promoting team-based models of care and supporting their implementation through research, policy briefs, and advocacy efforts (Table 3).

**Table 3.** Knowledge mobilization priorities and implementation tracks identified during Round Robin activity. Scores represent weighted priority rankings, with lower scores indicating higher priority. Track A includes initiatives considered feasible within the current funding cycle, and Track B represents longer-term priorities requiring additional development and resources.

| Content area | Knowledge mobilization strategy | Priority score | Feasibility track |
| --- | --- | --- | --- |
| Partnerships | Partner with BIC to identify community needs and co-develop targeted resources | 1.42 | B |
|  | Collaborate with federal agencies to mobilize knowledge across provinces | 2.17 | A |
|  | Partner with ABII to mobilize knowledge within Irish contexts | 4.91 | A |
|  | Collaborate with BIA and USA partners to adapt knowledge to different contexts | 5.33 | B |
| Centralized resource directory | Expand existing BIC and ABI Lab service directories to include additional Canadian services | 3.08 | A |
|  | Integrate existing directories into patient portals | 3.50 | A |
| Clinician education | Integrate concussion/TBI resources into *UpToDate* and similar platforms | 3.83 | A |
|  | Incorporate existing training opportunities into continuing medical education programs | 4.00 | B |
|  | Promote BIC landing page for healthcare professionals | 4.50 | A |
|  | Engage with clinician champions to promote TBI awareness and peer education | 6.00 | A |
| Integrated models of care | Promote team-based models of care across regions and specialties | 4.75 | B |
| Creative engagement | Develop creative outputs that amplify lived-experience perspectives | 4.83 | B |
|  | Apply for funds to reintroduce the *After the Crash* play across platforms | 5.50 | B |
| Future research | Use Canadian Community Health Survey data to investigate PROGRESS-Plus factors | 5.08 | B |
|  | Advocate for collection of social characteristic data in medical records | 5.08 | B |

**Abbreviations:** ABI, acquired brain injury; ABII, Acquired Brain Injury Ireland; BIA, Brain Injury Association; BIC, Brain Injury Canada; TBI, traumatic brain injury.

#### Evaluation and feedback

To evaluate the Round Robin activity and discussions, a standardized coding approach was applied to team member contributions to the discussion. Direct quotes were analyzed using the study-defined framework of FIDE, with multiple codes assigned where a quote reflected more than one principle (S11 File). Thematic analysis revealed variation in the extent to which FIDE principles were represented across discussion areas. Within the Information theme, five of eight team members addressed all four principles: feasibility, impact, development, and execution. In the Systems theme, one of four team members addressed all FIDE principles (S11 File). No stakeholders addressed all four principles within the Services and Benefits themes.

Within the more specific knowledge mobilization content areas, four team members addressed all FIDE principles when discussing centralized resources in Track A and future research, integrated care, and partnerships in Track B (S14 File).

#### Implementation and knowledge mobilization

As of May 15 2026, the publicly available report (49) had been accessed 1,058 times. The report includes targeted dissemination materials that were tailored to priority content areas for multiple stakeholder groups to enhance accessibility, uptake, and practical use of available services, including those within Brain Injury Canada (35) and the University of Toronto’s ABI Lab (48) service directories. These materials include resource repositories presented as interactive flipbooks, organized into two key domains: (1) education and training, and (2) wellness and support services. They also include bilingual (English and French) walkthrough videos of the Brain Injury Canada service directory to facilitate accessible, user-friendly navigation of relevant supports across stakeholder groups.

Peer-reviewed outputs have similarly demonstrated strong engagement and were led by trainees supported through the grant. The published protocol had been accessed 2,095 times since publication in July 2022 as of May 15, 2026 (19); meta-research on using ML to evaluate the relevance of social parameters for predicting cognitive outcomes following TBI (31) had been accessed 1,439 times within approximately a month and a half of publication (March 30 to May 15, 2026).

In addition, the research has been actively mobilized through presentations at local, national, and international conferences and scientific meetings, including the University of Toronto Rehabilitation Sciences Institute Research Day (2024), University Health Network’s Pride in Patient Engagement (PiPER) Research Day (2025), the Canadian Traumatic Brain Injury Research Consortium Trainee Day (2025), Canadian Consortium on Neurodegeneration in Aging Annual Conference (2025), the International Conference for Aging, Innovation, and Rehabilitation (2025), and the American Congress of Rehabilitation Medicine Annual Conferences (2024 and 2025) (40,41,50,51).

## Discussion

Our study provides a comprehensive, equity-oriented priority-setting analysis of social parameters affecting brain health in people with TBI, integrating evidence synthesis, stakeholder engagement, and structured consensus methods. Across multiple objectives, we identified key social parameters associated with brain health outcomes, mapped existing evidence using the PROGRESS-Plus framework, and translated these findings into actionable knowledge mobilization priorities. By combining empirical evidence with perspectives from people with lived experience, clinicians, researchers, and community leaders, our work illuminated a critical gap in the operationalization and consideration of social parameters within TBI research, policy, and practice concerning brain health. This gap underscores an urgent need for funding agencies, policymakers, and research institutions to support and act upon the proposed priority agenda to advance equity-oriented brain health.

The consistency in priorities across the countries and stakeholders’ groups suggests that social parameters affect brain health of people with TBI, despite differences in health systems and cultures. Shared priorities across stakeholders and continents – including access to benefits, level of formal education, and income – emerged as important for shaping brain health outcomes after TBI. Across stakeholder groups, specific priorities reflected unique lived and professional perspectives: patients and families emphasized trust in relationships; clinicians highlighted work status; and organizational leaders focused on workplace health and safety and age-related determinants. These priorities, summarized in the structured Round Robin ranking (Table 2), identified actionable items spanning system mobilization, service access, integration of resources into patient portals, capacity building through training, and development of team-based care models, demonstrating both global relevance and sensitivity to local context, providing a strong foundation for informing policy, service design, and equitable brain health care. However, translating these priorities into practice requires continued attention to how health and social systems can better align with the needs of people with TBI and their families.

There is currently significant consistency in the recommendations nationally and globally for social equity considerations in the prevention and care approach to brain health of people and communities (52). Many of these recommendations fall under the remit of government, but it is clear that to enable the change required for reduced TBI rates and its adverse brain health outcomes in the future, engagement from the full range of stakeholders is required, from people with lived experience and their families through community and brain injury organizations to the healthcare sector, researchers, and clinicians. In addition, it is clear that multiple institutional levels must play a role, including government through policy, legislation, and funding; civil society through advocacy, education, and community engagement; and the private sector through organization and delivery of evidence-based services, information management, funding, and accessibility (53). A successful approach to equity in brain health after TBI will require strong commitment and organizational leadership, and people with lived experience and community endorsement (17). Re-orienting clinically centered TBI activity environments alongside supporting and educating stakeholders’ communities will drive the culture change required to reduce and reverse adverse brain health outcomes after TBI. This is endorsed by the World Health Organization’s framework on meaningful engagement, which highlights practical guidelines, norms, and standards to transition from intention to action in operationalizing meaningful engagement (54). Evaluation techniques will need to continue to evolve to ensure implemented interventions are reflecting patients with lived experience and family needs, are equitable, and sustainable.

Policymakers should interpret these findings not as justification for maintaining current practices, but as evidence of minimum social conditions that must be prioritized in service design and resource allocation, which is needed to shape a more equity-driven and inclusive approach to policy-setting and implementation in neurological conditions including TBI (54). Accordingly, our recommendations (Figure 3) span four domains: strengthening information through improved integration and dissemination of PROGRESS-Plus considerations in knowledge mobilization, service directories, and patient-facing tools; embedding services that treat social determinants as core to brain health after TBI; advancing systems and infrastructure through coordinated interdisciplinary collaboration and investment in equitable workforce capacity; and improving benefits through comprehensive packages and multidisciplinary compensation models that support integrated, patient-centred care. At the governmental level, system optimization should follow a principle of social flexibility, ensuring that foundational social conditions are met while allowing adaptive variation across context-specific care systems.

### Strengths and limitations

Our research provides a novel and structured roadmap for future priority-setting studies integrating diverse methodological approaches, including systematic reviews, meta-research, surveys, and Round Robin consensus activities. This approach enables iterative evidence-based prioritization and consensus-building on key areas of knowledge mobilization. Our approach emphasized methodological transparency and inclusivity by documenting all stages of the process and avoiding the exclusion of priorities based on subjective judgments of relevance.

Outputs have been made available in an open-access repository (20) to increase the practical utility of findings for diverse stakeholder groups and accommodates a wide range of perspectives. Key strengths of this work include its cross-cultural scope, co-design with multiple stakeholder groups, and the utility of a mixed-methods approach. Survey participants were given opportunities to elaborate on their priorities and propose context-specific solutions, strengthening the interpretability and global applicability of the results. We are therefore confident that the most important social topics and themes were identified.

A limitation of the study that the numbers of respondents per stakeholder group were uneven, with a larger proportion of people with lived experience of TBI relative to other groups. This reflects successful engagement with the people and their needs but may also have influenced the balance of perspectives. To balance, we applied weighting to priorities based on number of participants in each group; however, the priority setting process is considered a consultancy process of a more qualitative nature, in which representativeness and saturation are more important than sample size. Given the consistency of findings across key stakeholders’ groups, we believe that saturation was reached.

A further limitation relates to the representation of stakeholders, particularly groups historically excluded from research (55–58). Despite targeted recruitment efforts and collaboration with global organizations representing people with TBI, and equity-focused leaders in brain health, survey respondents were predominantly White (Table 1). This limits the extent to which findings reflect the perspectives of people disproportionally affected by TBI and who carry a disproportional burden of adverse brain health outcomes (7). In addition, while we tried to distribute the survey globally, majority of the responses came from North America and Europe. This likely reflects representation on our team of Brain Injury Canada and the Global Brain Health Institute, Dublin headquarters, who circulated the survey among their networks. We took this limitation seriously and initiated multiple intentional and sustained engagement efforts with underrepresented groups in research and leadership coming from global regions for the co-creation of new grants.

In conclusion, in this research, we report the development of an evidence-based and stakeholder driven agenda situated around importance of consideration of social parameters in research, policy, and practice. We highlighted both shared and stakeholder-specific priorities. The resulting agenda sets a solid foundation for guiding future research, health system responses, and policy development. Our multidisciplinary team has close ties to community of diverse backgrounds, offering valuable opportunities to disseminate findings to knowledge users and decision-makers across the globe, through multiple consortia, professional organizations, and networks, including the North American Brain Injury Society, Canadian Traumatic Brain Injury Consortium, American Congress of Rehabilitation Medicine, and the Global Brain Health Institute, each of which has its own knowledge brokers to facilitate knowledge translation and exchange support to reach a global audience. We anticipate that our research will add significant value to the field of TBI by promoting equity-transformative scientific advancements in knowledge synthesis, policy, and practice to enhance access to knowledge for all.

## Data Availability

Data used in this study can be accessed in the supporting files and OSF data repository: https://osf.io/5sjvx

## Author contributions

Conceptualization: Sara Hanafy, Esme Fuller-Thomson, Michelle McDonald,

Angela Colantonio, DaíthíCee, Gráinne McGettrick, Brian Lawlor, Tatyana Mollayeva.

Data curation: Thaisa Tylinski Sant’Ana, Tatyana Mollayeva.

Formal analysis: Thaisa Tylinski Sant’Ana, Urooba Shaikh, Rachel Spouge, Tatyana Mollayeva.

Funding acquisition: Tatyana Mollayeva.

Investigation: Thaisa Tylinski Sant’Ana, Urooba Shaikh, Rachel Spouge, Sara Hanafy, Esme Fuller-Thomson, Michelle McDonald, Angela Colantonio, DaíthíCee, Gráinne McGettrick, Brian Lawlor, Tatyana Mollayeva.

Methodology: Thaisa Tylinski Sant’Ana, Tatyana Mollayeva. Project administration: Thaisa Tylinski Sant’Ana, Urooba Shaikh

Resources: Michelle McDonald, DaíthíCee, Gráinne McGettrick, Brian Lawlor, Tatyana Mollayeva.

Supervision: Tatyana Mollayeva.

Visualization: Urooba Shaikh, Tatyana Mollayeva.

Writing – original draft: Urooba Shaikh, Rachel Spouge, Tatyana Mollayeva.

Writing – review & editing: Thaisa Tylinski Sant’Ana, Sara Hanafy, Esme Fuller-Thomson, Michelle McDonald, Angela Colantonio, DaíthíCee, Gráinne McGettrick, Brian Lawlor. All authors contributed to the research, as well as read and approved the final manuscript.

## Acknowledgements

This research was conducted, reported, and reviewed by team members chosen for their diverse perspectives and interdisciplinary expertise. The team would like to express its gratitude to the many people and families dealing with TBI and the researchers, clinicians, and leaders of the organizations that represent people with TBI who generously lent their time and insights to complete the survey that allowed us to see TBI truly in all its dimensions, and for sharing their concerns in a health care system that lacking their social needs in TBI care. The survey results led informed priority setting Round Robin and knowledge mobilization activities and led the research team to inescapably event the priorities of people with lived experience in knowledge mobilization. We owe a deep debt of gratitude to Daithi Cee. Their thoughtful feedback, reflections, and contributions throughout the research helped shape and advance the team’s vision for diversity and inclusion. We are especially grateful for their generosity in contributions across countries, time zones, and disciplinary boundaries, and for their extraordinary humanity and lifelong advocacy for the LGBTQ2+ community in the face of adversity. The team is also grateful for the contributions of trainees of the BRIDGE Lab, University Health Network (59): Ashlee Kim, Chuxi Pan, Mursal Jahed, Daniel Phan, Jingwen Xu, Yupeng Yao, Sophia Karim, Alexandra Cuyugan, Dian Zhang, Noshin Haque, Christel Costa Tiago, and Darren Tang, for their crucial contributions to the advancement of social equity research, their endorsement of the PROGRESS-Plus framework in the vision of improvement of brain health for all, and their extended knowledge mobilization activities.

## Our vision and values

Setting priorities involved making choices, and those choices referred to underlying values of the team: Authenticity, Fairness, Openness, Respect, Courage, and Empathy. We set priorities in an ethically appropriate, evidence-based way, identified evidence gaps regarding social parameters considerations in TBI globally and considered a range of contexts and relevant stakeholders (e.g., people with TBI, their family/friends, healthcare providers, researchers, and staff of TBI organizations). We provided actionable recommendations for a solid strategy to inform better policies and practices for people with TBI, in the context of brain health outcomes.

## Supporting information captions

**S1 File.** Ethics approval from University Health Network, Toronto, Canada

**S2 File.** Team members’ affiliations, expertise, and roles within the research project.

**S3 File.** Participant information sheet distributed with survey.

**S4 File.** Poll distributed to team members for selection of working definition of brain health and weighted results.

**S5 File.** Social parameters relevant to brain health, structured using the PROGRESS-Plus framework.

**S6 File.** Contents of preliminary survey distributed to team members for review.

**S7 File.** Contents of final version of survey distributed to stakeholders globally.

**S8 File.** Descriptive tables of number of respondents who selected the PROGRESS-Plus category as their top priority.

**S9 File.** Results of thematic analysis of responses to the question “What resources would help you better understand the links between brain health and social factors? Your response can refer to resources for people with brain injury, their families, and professionals in the field.”

**S10 File.** Team members’ selections of top two knowledge translation categories.

**S11 File.** Summary of key themes that emerged from team discussions during Round Robin activity.

**S12 File.** Round Robin activity agenda.

**S13 File.** Round Robin Activity results.

**S14 File.** Content areas and principles discussed in Round Robin Activity Round 2. Green indicates where all FIDE principles were discussed in relation to the content area.

